# Germline mutation rates in young adults predict longevity and reproductive lifespan

**DOI:** 10.1101/19004184

**Authors:** Richard M. Cawthon, Huong D. Meeks, Thomas A. Sasani, Ken R. Smith, Richard A. Kerber, Elizabeth O’Brien, Lisa Baird, Melissa M. Dixon, Andreas P. Peiffer, Mark F. Leppert, Aaron R. Quinlan, Lynn B. Jorde

**Author notes:** Corresponding author: R.M.C. H.D.M. and T.A.S. contributed equally to this article.

## Abstract

**BACKGROUND:** Analysis of sequenced genomes from large three-generation families allows *de novo* mutations identified in Generation II individuals to be attributed to each of their parents’ germlines in Generation I. Because germline mutations increase with age, we hypothesized that they directly limit the duration of childbearing in women, and if correlated with mutation accumulation in somatic tissues, also reflect systemic aging in both sexes. Here we test whether the germline mutation rates of Generation I individuals when they were young adults predict their remaining survival, as well as the women’s reproductive lifespans.

**METHODS:** Germline autosomal mutation counts in 122 Generation I individuals (61 women, 61 men) from 41 three-generation Utah CEPH families were converted to germline mutation rates by normalizing each subject’s number of mutations to the callable portion of their genome. Age at death, cause of death, all-site cancer incidence, and reproductive histories were provided by the Utah Population Database, Cancer Registry, and Utah Genetic Reference Project. Fertility analyses were restricted to the 53 women whose age at last birth (ALB) was at least 30 years, the approximate age when the decline in female fertility begins. Cox proportional hazard regression models were used to test the association of age-adjusted mutation rates (AAMRs) with aging-related outcomes. Linear regression analysis was used to estimate the age when adult germline mutation accumulation rates are established.

**FINDINGS:** Quartiles of increasing AAMRs were associated with increasing all-cause mortality rates in both sexes combined (test for trend, p=0.009); subjects in the top quartile of AAMRs experienced more than twice the mortality of bottom quartile subjects (hazard ratio [HR], 2.07; 95% confidence interval [CI], 1.21-3.56; p=0.008; median survival difference = 4.7 years). Women with higher AAMRs had significantly fewer live births and a younger ALB. The analyses also indicate that adult germline mutation accumulation rates are established in adolescence, and that later menarche in women may delay mutation accumulation.

**INTERPRETATION:** Parental-age-adjusted germline mutation rates in healthy young adults may provide a measure of both reproductive and systemic aging. Puberty may induce the establishment of adult mutation accumulation rates, just when DNA repair genes’ expression levels are known to begin their lifelong decline.

**FUNDING:** NIH R01AG038797 and R21AG054962 (to R.M.C.); University of Utah Program in Personalized Health (to H.D.M.); NIH T32GM007464 (to T.A.S.); NIH R01AG022095 (to K.R.S.); NIH R01HG006693, R01HG009141, and R01GM124355 (to A.R.Q.); NIH GM118335 and GM059290 (to L.B.J.); NIH P30CA2014 (to the Utah Population Database, a.k.a. the UPDB); National Center for Research Resources Public Health Services grant M01RR00064 (to the Huntsman General Clinical Research Center, University of Utah); National Center for Advancing Translational Sciences NIH grant UL1TR002538 (to the University of Utah’s Center for Clinical and Translational Science); Howard Hughes Medical Institute funding (to Ray White); gifts from the W.M. Keck Foundation (to Stephen M. Prescott and M.F.L.) and from the George S. and Delores Doré Eccles Foundation (to the University of Utah) that supported the Utah Genetic Reference Project (UGRP). Sequencing of the CEPH samples was funded by the Utah Genome Project, the George S. and Dolores Doré Eccles Foundation, and the H.A. and Edna Benning Foundation. We thank the Pedigree and Population Resource of the Huntsman Cancer Institute, University of Utah (funded in part by the Huntsman Cancer Foundation) for its role in the ongoing collection, maintenance and support of the UPDB.

## INTRODUCTION

The somatic mutation theory of aging^1^ proposes that somatic mutations accumulate throughout life, resulting in apoptosis, cellular senescence, tumorigenesis, or other cellular pathologies, followed by tissue dysfunction, chronic disease, and death. DNA damage is continuous,^2^ and while most of it is repaired, several classes of DNA damage are known to accumulate with age in both sexes,^3-6^ though at higher rates in men.^7^ Serum levels of insulin-like growth factor I (IGF-I) peak during puberty,^8^ suppressing the FOXO transcription factors and the DNA repair genes that FOXO upregulates.^9^ DNA repair systems continue to decline throughout adult life.^10^ Furthermore, developmental deficiency of the GH/IGF-1 axis in dwarf mice prevents the normal decline in DNA repair of adulthood and significantly extends lifespan.^11^ These observations fit well with the evolutionary biology principle that the force of natural selection to maintain robust health begins to decline once the reproductive phase of life is attained.^12,13^

Several monogenic segmental progeroid syndromes in humans are known to increase mutation accumulation rates and shorten the lifespan.^14^ However, no studies have yet tested whether inter-individual differences in either somatic or germline mutation accumulation rates in healthy young adults predict differences in remaining lifespan, as would be expected if mutation accumulation contributes significantly to aging. Fertility in healthy women declines after age 30;^15^ age at natural menopause varies from 40-60 years and is positively associated with longer post-reproductive lifespans;^16^ and women with older ALB and their siblings enjoy increased longevity.^17^ Whether variation between healthy women in the decline and end of fertility may be attributable, at least in part, to differences in germline mutation accumulation rates is unknown. Similarly, while somatic mutations are known contributors to tumorigenesis,^18^ a connection between mutation accumulation rates in healthy young adults and cancer risks has not yet been established.

Recently, we analyzed^19^ whole genome sequencing (WGS) data from 41 three-generation Utah CEPH (Centre d’Etude du Polymorphisme Humain) families^20,21^ and identified mutations that arose in the germ cells of the Generation I (oldest) individuals and were transmitted to their Generation II offspring. Here we test whether parental-age-adjusted germline autosomal mutation rates in Generation I are associated with two clinically important life history traits in those same individuals: lifespan in both sexes and the duration of childbearing in women, as would be expected if germline mutation accumulation reflects the rate of both systemic and reproductive aging. We also investigate the hypothesis that puberty initiates the establishment of adult germline mutation accumulation rates following a prepubertal quiescent period when mutation burdens may be plateaued.

## METHODS

### HUMAN SUBJECTS

In the early 1980s, the 46 three-generation Utah CEPH families were contacted and enrolled in a project to build the first comprehensive human genetic linkage map.^20,21^ Each Utah CEPH family consists of 4–16 siblings in the youngest generation (Generation III), their two parents (Generation II), and two to four grandparents (Generation I), as shown in Figure S1 of the Appendix. Here we study 122 Generation I individuals (61 women and 61 men; demographic characteristics presented in Table S1) from 41 of the families, as well as 40 Generation II parental couples and their 350 Generation III offspring. DNA was extracted from blood samples collected in the early 1980s and/or early 2000s. All participants provided written informed consent, and all studies were conducted with University of Utah Institutional Review Board approval.

### AGE-ADJUSTED GERMLINE MUTATION RATES

*De novo* mutations in the germ cells of parents can be found by WGS of DNA extracted from somatic tissue (e.g. blood samples) from parents and offspring, identifying high-confidence sequence changes in the offspring not present in either parent, and attributing each mutation to the parental germline in which it arose.^19,22,23^ The number of germline mutations increases with parental age in both sexes, with higher absolute levels and higher rates of accumulation in males, and mutation counts varying more than two-fold between age-matched individuals of the same sex.^19,23,24^

WGS of blood DNA from 603 individuals from 41 three-generation families, identification of autosomal *de novo* mutations (single base substitutions and insertions and deletions of length 10 basepairs or less) in Generation II, and specific attribution of each mutation to the germline of a Generation I individual is described by Sasani and colleagues.^19^ Instead of simple counts of Generation I germline mutations, we analyzed germline mutation *rates*, obtained by dividing the number of mutations by the callable portion of the subject’s genome (number of autosomal mutations / number of diploid autosomal callable basepairs), to adjust for minor differences between subjects in the portion of the genome that met our requirements for validating mutations (Sasani et al.,^19^ pp. 14-16). The Generation I germline autosomal mutation rates that were the basis for all analyses presented here are plotted against parental age in Figure S2.

Because germline mutation counts increase with age in both sexes, we regressed mutation rates on parental age using a general linear model and used the resulting residuals to represent age-adjusted mutation rates (AAMRs). AAMRs treated either as a continuous or as a categorical variable were tested for their associations with aging-related outcomes. The median ages of subjects in each quartile and tertile of AAMRs are given in Table S2.

### OUTCOMES

Generation I subjects were linked to the Utah Population Database (UPDB), a large and comprehensive resource of linked population-based information for demographic, genetic, and epidemiological studies (https://uofuhealth.utah.edu/huntsman/utah-population-database/acknowledging-updb.php). The UPDB is a dynamic genealogical and medical database that receives annual updates of Utah birth, death, and health records. This study received approval from the University of Utah’s Resource for Genetic and Epidemiologic Research (RGE) and Institutional Review Board (IRB). Mortality was ascertained based on Utah death certificates linked to the UPDB. Causes of death were available in International Classification of Diseases (ICD) codes version 9-10 and aggregated into larger categories representing the leading causes of deaths. Cancer incidence records were drawn from the Utah Cancer Registry (https://uofuhealth.utah.edu/utah-cancer-registry/). Fertility in Generation I women was assessed by parity (number of live births) and ALB, both derived from the UPDB. Self-reported age at menarche (answer to the question, “At what age did your menstrual periods start [age in years]?”) was provided by the Utah Genetic Reference Project (UGRP).

### STATISTICAL ANALYSIS

Full sample and sex-specific Cox proportional hazard models with adjustments for subject’s birth year were used to estimate the effects of AAMRs on mortality and cancer risks, expressed as hazard rate ratios (HR) in Generation I individuals. Results for both sexes combined were adjusted for sex. Time was measured in years from the Generation I individual’s parental age at the birth of the index child to time of death (n=120) or last known living dates up to 2018 (n=2). Cause-specific mortality was analyzed by fitting cause-specific hazard regression models with Cox regression, treating failures from the cause of death of interest as events and failure from other causes of deaths or those still living as right-censored.^25^ For cancer incidence analyses, subjects were censored at age at death or age at last follow-up, whichever occurred first.

Poisson regression models were used to assess the effect of AAMRs on the number of live births to Generation I women. Logistic regression models were used to assess the effect of AAMRs on ALB where ALB was treated as a categorical variable (i.e. <25^th^ percentile). All fertility analyses in these Generation I women were adjusted for subject’s birth year and only included women with an ALB ≥ 30 years, since cessation of childbearing prior to age 30 is unlikely due to reproductive aging.^15^

The association of AAMRs with age at menarche in the 20 Generation I women for whom menarcheal age was available was tested by a Pearson correlation (r) analysis, using 5000 bootstrapped samples, given the small sample size. The significance of the statistic (r in this case) is an empirical (bootstrapped) p value and confidence intervals. The difference in mean age at menarche between women with AAMRs ≥ 50^th^ percentile vs. women below the 50^th^ percentile was also analyzed.

## RESULTS

### CHARACTERISTICS OF THE SUBJECTS

In developing genetic linkage maps, the large sibship sizes of the Utah CEPH families allowed the segregation of genetic markers to be replicated in informative families, and the inclusion of grandparents helped in assigning alleles to maternal and paternal chromosomes (phasing).^20,21^ The families were not selected for any disease, but for large sibship sizes. Selecting for large sibships may select somewhat for higher than average fertility, and selecting for living grandparents may select somewhat for higher than average lifespan; however, large sibships are common in Utah, and more than half of the grandparents were younger than age 72 at the time of the initial enrollment. Therefore, these families are unlikely to be strongly enriched for factors contributing to longer reproductive lifespans and longer life. Furthermore, since the same selection criteria were applied across all collected families, these criteria should not introduce any biases for the current study. Finally, nearly all the Utah CEPH grandparents are of Northern European descent.

### SURVIVAL ANALYSES

Associations of AAMRs with all-cause mortality, cardiovascular disease (CVD) mortality, and non-CVD mortality were analyzed in both sexes combined and in each sex separately (Table 1). CVD mortality includes deaths from heart disease, stroke, and hypertension. This approach revealed differences between men and women in the causes of mortality most strongly associated with increasing germline mutation rates.

**Table 1:** Effect of germline mutation rates on mortality in 122 Generation I individuals.

|  | Age-adjusted<br>germline<br>mutation rates | All-cause mortality |  | CVD mortality |  | Non-CVD mortality |  |
| --- | --- | --- | --- | --- | --- | --- | --- |
|  |  | HR (95% CI) | p | HR (95% CI) | p | HR (95% CI) | p |
| Both sexes | Continuous | 1.28 (1.05, 1.56) | **0.015** | 1.14 (0.87, 1.49) | 0.355 | 1.49 (1.12, 2.00) | **0.007** |
|  | 25<sup>th</sup>-50<sup>th</sup> percentile | 1.51 (0.88, 2.59) | 0.136 | 1.67 (0.79, 3.50) | 0.177 | 1.41 (0.63, 3.14) | 0.406 |
|  | >50<sup>th</sup>-75<sup>th</sup> percentile | 1.59 (0.94, 2.70) | 0.087 | 1.29 (0.57, 2.91) | 0.545 | 2.07 (1.01, 4.24) | **0.047** |
|  | > 75<sup>th</sup> percentile | 2.07 (1.21, 3.56) | **0.008** | 1.28 (0.56, 2.93) | 0.559 | 3.24 (1.57, 6.68) | **0.001** |
|  | Trend test | 1.25 (1.06, 1.48) | **0.009** | 1.05 (0.82, 1.34) | 0.694 | 1.48 (1.18, 1.87) | **0.001** |
| Males | Continuous | 1.40 (1.04, 1.87) | **0.025** | 1.39 (0.94, 2.07) | 0.101 | 1.45 (0.94, 2.22) | 0.092 |
|  | 25<sup>th</sup>-50<sup>th</sup> percentile | 1.24 (0.58, 2.65) | 0.578 | 1.62 (0.55, 4.76) | 0.383 | 1.11 (0.37, 3.37) | 0.849 |
|  | >50<sup>th</sup>-75<sup>th</sup> percentile | 1.80 (0.85, 3.83) | 0.125 | 2.64 (0.86, 8.17) | 0.091 | 1.45 (0.51, 4.14) | 0.483 |
|  | > 75<sup>th</sup> percentile | 2.21 (1.03, 4.76) | **0.043** | 2.39 (0.77, 7.46) | 0.132 | 2.27 (0.79, 6.51) | 0.126 |
|  | Trend test | 1.32 (1.03, 1.68) | **0.026** | 1.35 (0.96, 1.90) | 0.084 | 1.31 (0.93, 1.85) | 0.122 |
| Females | Continuous | 1.22 (0.93, 1.59) | 0.158 | 0.74 (0.49, 1.12) | 0.152 | 1.76 (1.24, 2.51) | **0.002** |
|  | 25<sup>th</sup>-50<sup>th</sup> percentile | 2.04 (0.91, 4.59) | 0.085 | 1.63 (0.56, 4.71) | 0.368 | 2.17 (0.61, 7.64) | 0.230 |
|  | >50<sup>th</sup>-75<sup>th</sup> percentile | 1.44 (0.67, 3.10) | 0.350 | 0.43 (0.11, 1.69) | 0.225 | 2.91 (1.06, 8.03) | **0.039** |
|  | > 75<sup>th</sup> percentile | 1.97 (0.88, 4.38) | 0.097 | 0.41 (0.10, 1.60) | 0.199 | 5.16 (1.79, 14.93) | **0.002** |
|  | Trend test | 1.18 (0.93, 1.50) | 0.180 | 0.70 (0.47, 1.04) | 0.081 | 1.68 (1.21, 2.33) | **0.002** |
| Associations with mortality of age-adjusted mutation rates (AAMRs) treated as a continuous or categorical variable were analyzed in both sexes combined and in each sex separately. In each section (Both sexes, Males, and Females) the first row (Continuous) presents the effects on the Hazard Ratio (HR) of a one standard deviation increase in AAMRs. The second, third, and fourth rows present the mortality risks for subjects with increasing quartiles of AAMRs, expressed relative to the mortality risks for the lowest quartile (<25<sup>th</sup> percentile). The thresholds for the AAMRs are Males: 25% = - 1.2385992, 50% = - 0.1078028, 75% = 1.2402645; Females: 25% = - 0.61610326, 50% = - 0.09220471, 75% = 0.41736567. These thresholds were calculated using all 122 Generation I subjects. CI = Confidence Interval. |  |  |  |  |  |  |  |

After adjusting for birth year and sex in Cox proportional hazard regression models, in both sexes combined a one standard deviation increase in AAMRs was associated with higher all-cause mortality (hazard ratio [HR], 1.28; 95% confidence interval [CI], 1.05-1.56; p=0.015), and higher non-CVD mortality (HR, 1.49; 95% CI, 1.12-2.00; p=0.007), but not associated with CVD mortality.

Among men a one standard deviation increase in AAMRs was significantly associated with higher all-cause mortality (HR, 1.40; 95% CI, 1.04-1.87; p=0.025), but not associated with either non-CVD or CVD mortality.

Among women a one standard deviation increase in AAMRs was associated with non-CVD mortality (HR, 1.76; 95% CI, 1.24-2.51; p=0.002), but not associated with either all-cause mortality or CVD mortality.

All categorical comparisons in Table 1 used < 25^th^ percentile of AAMRs as the reference category. Similar to the above-noted associations between continuous AAMRs and mortality risk, tests for trend across increasing quartiles of AAMRs were statistically significant. The categorical analyses suggest that in men, both CVD mortality and non-CVD mortality contribute to the significant association of AAMRs with all-cause mortality. In contrast, in women the association of AAMRs with mortality appears to be driven almost entirely by their association with non-CVD mortality. (When mutation rates were adjusted for parental age by simply dividing each subject’s mutation rate by their parental age, similar, though less robust associations with all-cause mortality risks were observed: Table S3.)

The higher germline mutation counts and higher rates of accumulation of germline mutations in men vs. women have been attributed to the male, but not the female, germline undergoing rounds of genome copying and cell division throughout adulthood that generate replication errors; in addition there appear to be higher rates of unrepaired DNA damage (independent of genome replication) in male vs. female germlines.^24^ How these sex differences in germline mutation dynamics may relate to the different patterns of associated cause-specific mortality in men vs. women remains to be elucidated.

Figure 1 presents adjusted survival curves by quartile of AAMRs. The median survival advantage for all-cause mortality in the men and women combined analysis, for those with mutation rates < 25th percentile vs. mutation rates > 75th percentile, was approximately 4.7 years. For male all-cause mortality, the median survival advantage for bottom quartile vs. top quartile was approximately 6 years. For female non-CVD mortality, the median survival advantage for bottom quartile vs. top quartile was approximately 8 years.

**Figure 1:**
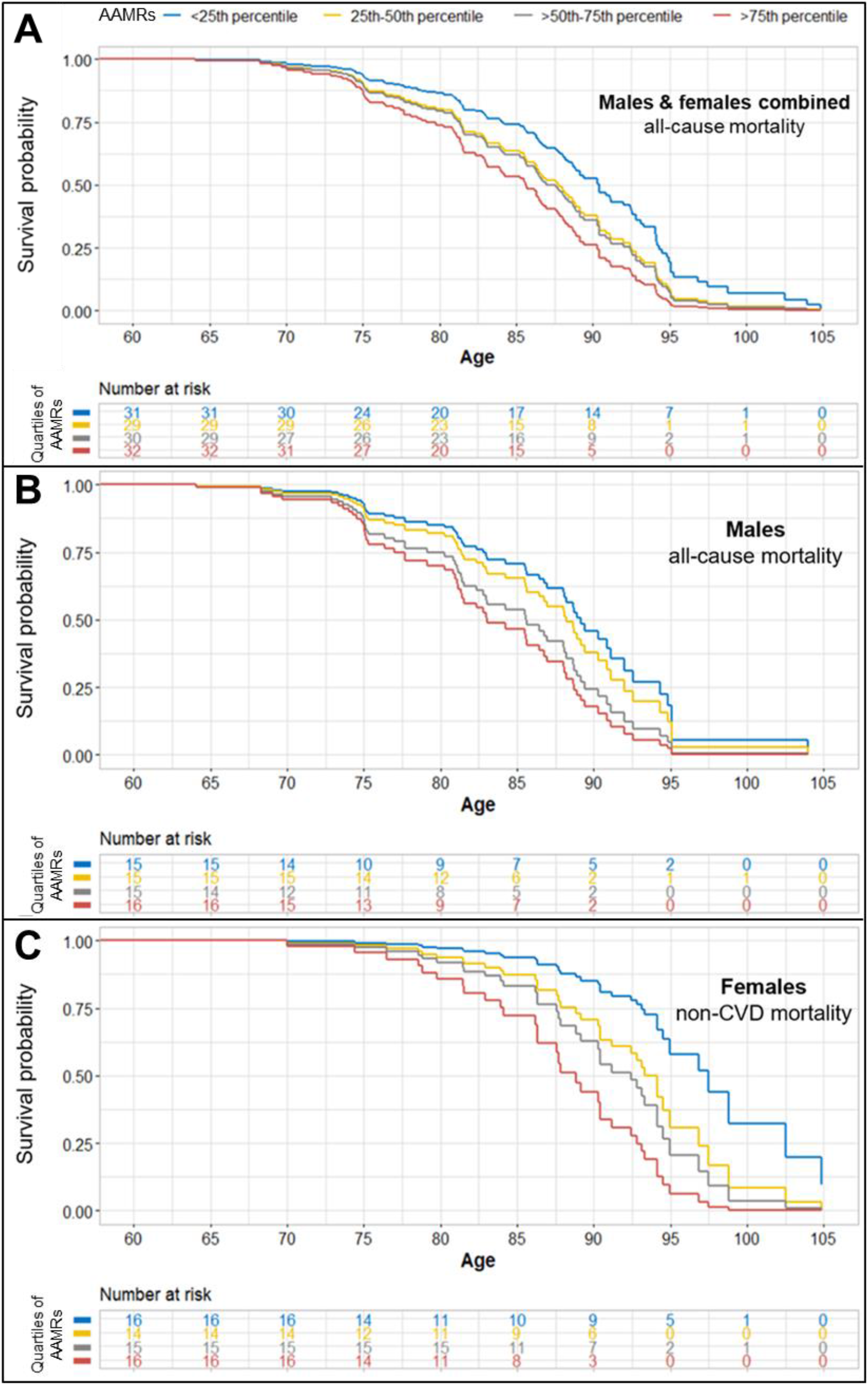
Predicted survival curves by quartiles of age-adjusted germline mutation rates. Parental age and birth year were fixed to their median values (25 years and 1912, respectively) based on the fitted model in Table 1. Panel A: both sexes combined, all-cause mortality; Panel B: males only, all-cause mortality; Panel C: females only, non-cardiovascular disease (non-CVD) mortality. AAMRs: age-adjusted mutation rates.

Germline mutation rates in adults are known to be much lower than somatic mutation rates.^26^ However, our results are consistent with a model whereby inter-individual differences in germline mutation rates may correlate with inter-individual differences in the rates of accumulation of somatic mutations that contribute to the development of multiple aging-related lethal diseases.

### CANCER INCIDENCE

In the set of 122 Generation I individuals, there were 16 women and 18 men who received at least one cancer diagnosis in their lifetimes. We sought to test the hypothesis that in the full cohort of 122 subjects, lower AAMRs would be associated with lower age-specific cancer risks (Table S4). Associations were tested with AAMRs treated as a continuous variable and also as a categorical variable where cancer risks in the higher tertiles of mutation rates are compared to cancer risk in the lowest tertile. No associations of germline mutation rates with cancer risk were found. Similar associations tested with larger cohorts are needed to further investigate this hypothesis.

### FERTILITY OF WOMEN

We hypothesized that germline mutation accumulation may contribute to oocyte atresia, lower rates of fertilization, higher rates of miscarriage, and/or earlier menopause, and consequently, shorter reproductive lifespans. Therefore, we tested whether women with higher rather than lower AAMRs gave birth to fewer children, and had a younger ALB (Table 2). The thresholds for mutation rates and the threshold for ALB < 25^th^ percentile were calculated using the 53 Generation I women with age at last birth ≥ 30. Among these women, those in the top two thirds of AAMRs had fewer live births than those in the bottom third (p=0.018), and higher mutation rates were significantly associated (p=0.036) with a younger ALB (< 25th percentile, 34.8 years).

**Table 2:** Effects of germline mutation rates on reproductive lifespan in 53 Generation I women with ALB ≥30 years.

| Age-adjusted<br>germline<br>mutation rates | Number of live births |  |  |  | Age at last birth<br><25<sup>th</sup> percentile |  |
| --- | --- | --- | --- | --- | --- | --- |
|  | Est | SE | Z | p | RR (95% CI) | p |
| Continuous | -0.12 | 0.08 | -1.63 | 0.104 | 2.12 (1.05, 4.26) | 0.036 |
| ≥ 33<sup>rd</sup> percentile | -0.27 | 0.11 | -2.36 | 0.018 | 4.27 (0.81, 22.41) | 0.086 |
| Effects of AAMRs as a continuous or categorical variable on the number of live births and ALB for the 53 Generation I women with ALB ≥30 years. For the categorical analyses, women in the top two thirds of AAMRs were compared to women in the bottom third. Poisson regression was used to assess the effect of AAMRs on the number of live births, additionally adjusted for birth year. The number of live births decreased by 8.73% for each standard deviation increase in the AAMRs (p = 0.104). Logistic regression models were used to assess the association of AAMRs with ALB, additionally adjusted for birth year and whether the subject had any children with missing birth dates in UPDB. The risk of the ALB being below the 25<sup>th</sup> percentile increased 2.12 times for every standard deviation increase in the AAMRs (95% CI 1.05-4.26), p = 0.036). The 33<sup>rd</sup> percentile cut point for AAMRs was - 0.46200203. The 25<sup>th</sup> percentile cut point for age at last birth was 34.8 years. |  |  |  |  |  |  |

### WHEN ARE THE GERMLINE MUTATION ACCUMULATION RATES OF ADULTHOOD ESTABLISHED?

Our data suggest that germline mutation accumulation rates in young adults may be a measure of the rate of aging. Sasani et al.^19^ demonstrated (in their Figure 3c and 3d) a 3+ fold range of germline mutation accumulation rates in 40 CEPH Generation II parental couples (primarily reflecting the levels and rates of accumulation of mutations in the fathers’ germlines). Taken together these data suggest that the rate of aging may vary 3-fold between young adults. Recent studies of inter-individual variation in the pace of aging in young adults, based on measures of physical performance, physiological functioning, cellular and biochemical markers, and gum health also found approximately 3-fold variation between individuals.^27,28^

When during human development are differences between individuals in the rate of aging established? Published models suggest that germline mutation counts may be plateaued prepubertally.^29^ The risk of dying is also plateaued and at its nadir during the prepubertal years.^30^ Together, these observations support the hypothesis that aging begins at or soon after puberty, due to a decline in the force of natural selection to maintain robust health once the reproductive phase of life is attained.^12,13^

To further investigate this hypothesis, we first estimated when the germline mutation accumulation rates of adulthood are established, by plotting germline mutation rates (x axis) vs. parental age (y axis) for our 122 Generation I subjects (Figure 2). The y intercepts are the ages when the mutation rates would equal zero. We interpret the y intercepts as approximate lower bounds for the ages when the observed mutation accumulation rates began: ∼18 years for females and ∼14 years for males. This timing suggests that puberty may indeed have a causal role in establishing the mutation accumulation rates of adulthood.

**Figure 2.**
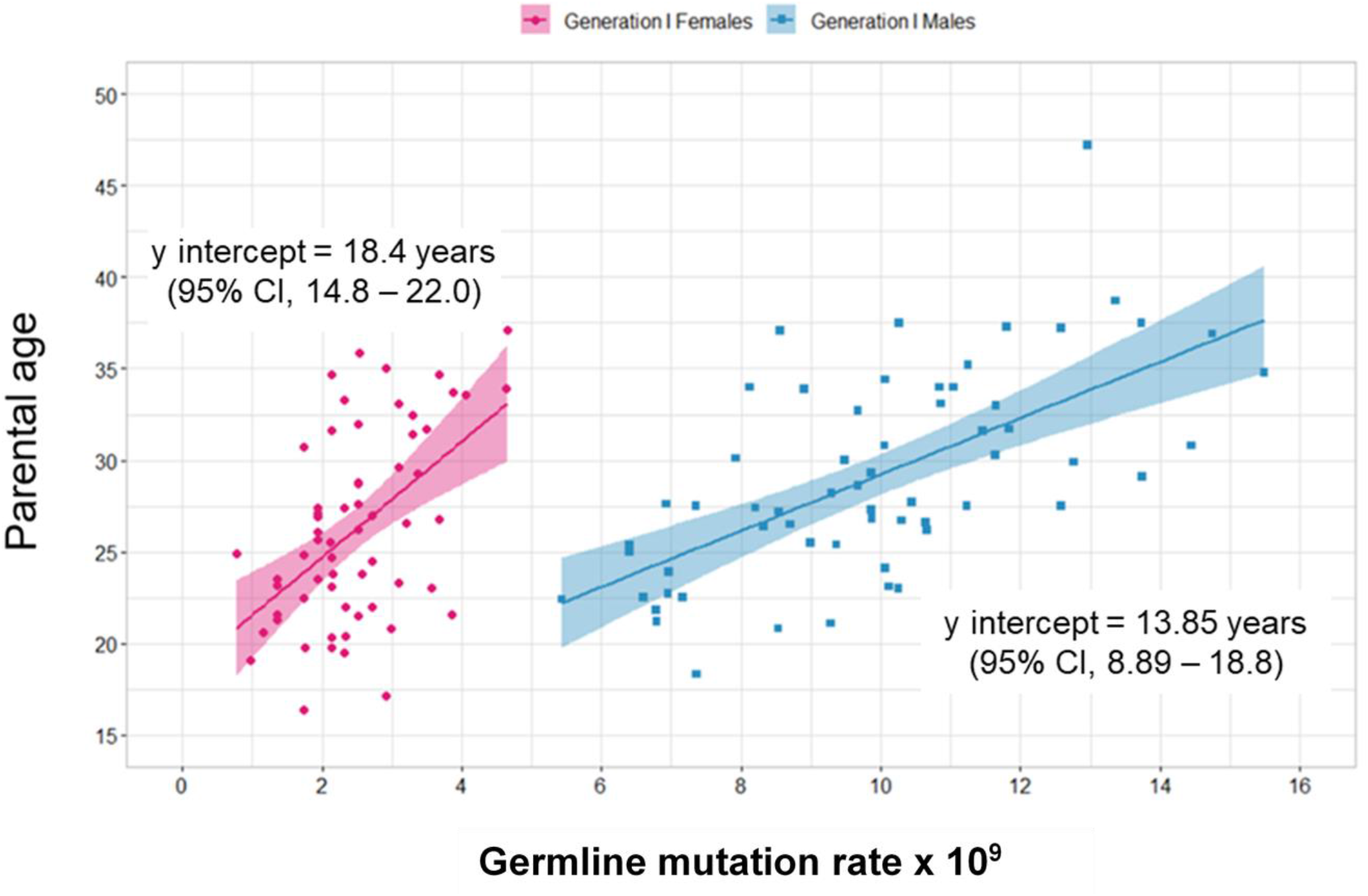
Estimating the age when adult germline mutation accumulation rates are established. Cross-sectional data for germline autosomal mutations in 122 Utah CEPH Generation I individuals, based on *de novo* mutations discovered in one Generation II offspring per Generation I parental couple. The y intercepts of the linear regression lines, when mutation counts would be zero, provide approximate lower bounds for the age when the observed mutation accumulation rates (slopes of the regression lines) were established: about 18 years for Generation I women and 14 years for Generation I men. Germline mutation rate = (#germline autosomal mutations) / (#diploid autosomal callable basepairs).

**Figure 3.**
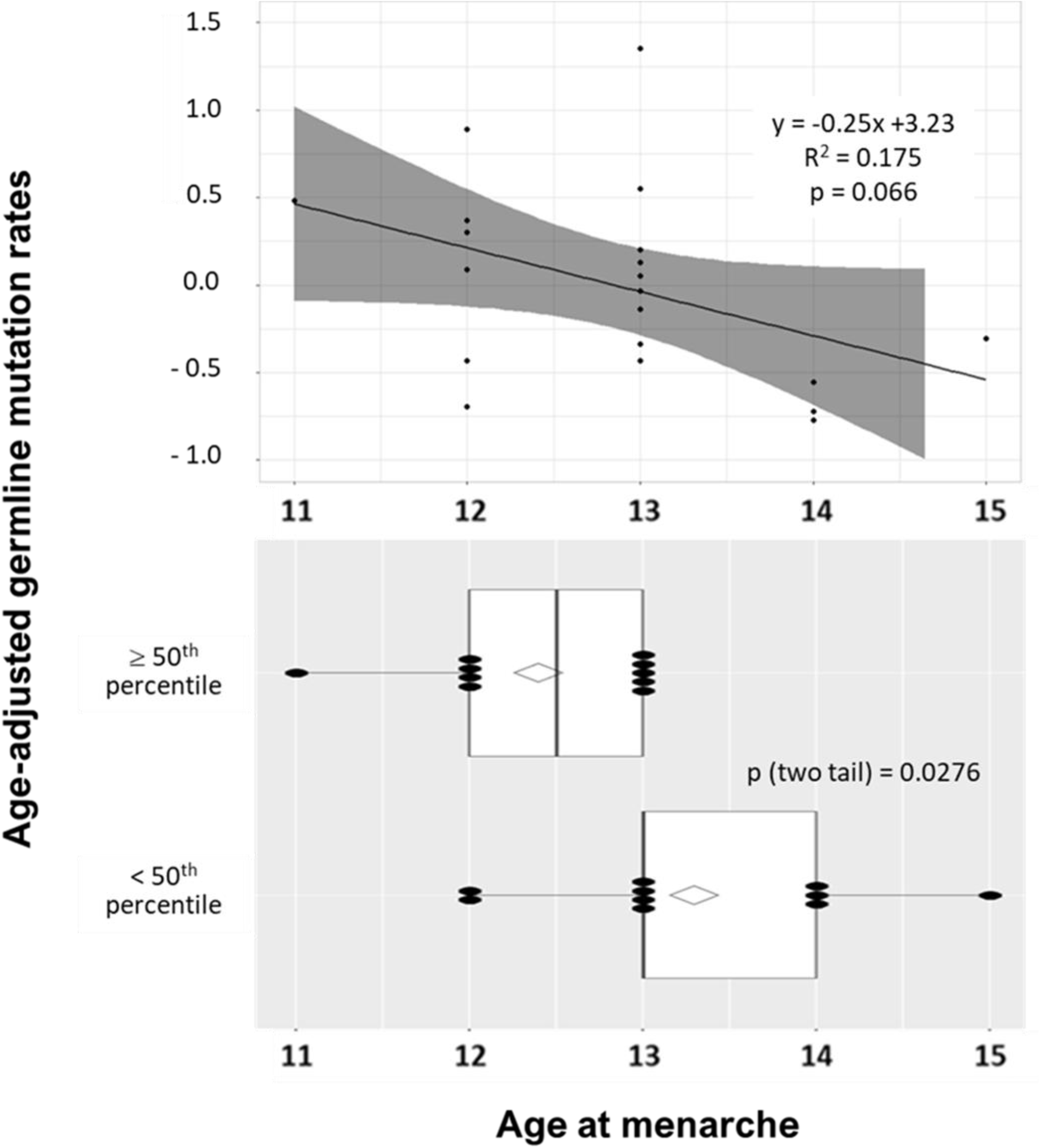
Effects of age at menarche on germline mutation rates in 20 Generation I women. Top panel: linear regression of AAMR by age at menarche. Bottom panel: box plot of age at menarche by two categories of AAMRs, ≥ 50th percentile (≥ 0.0095) and < 50th percentile (< 0.0095). Diamonds represent the mean age of menarche (12.4 years for AAMRs ≥ 50th percentile and 13.3 years for AAMRs < 50th percentile). Dots represent the ages of menarche. Using the t-test to test differences between means of age at menarche for AAMRs < 50th percentile vs. ≥ 50th percentile, p-value = 0.0276.

We also tested another prediction of the hypothesis, that later onset of puberty should be accompanied by delays in mutation accumulation and lower age-adjusted mutation rates. Age at menarche data were available for 20 of the Generation I women. We found (Figure 3) that older ages at menarche were associated with lower AAMRs, and that women below vs. above the median mutation rate had a higher mean age at menarche (two-tailed t-test, p=0.028). When tested by a Pearson correlation (r) analysis, the association of AAMRs with age at menarche was also significant (r = −0.418, p=0.021).

Finally, since the range of onset of puberty is about five years,^31^ the hypothesis further predicts that adults with equivalent mutation accumulation rates should vary about five years in the age at which they acquire the same number of mutations, a prediction supported by our longitudinal data on germline mutation accumulation in parental couples (Figure S3).

Taken together, these results from both sexes suggest a model whereby puberty induces germline mutation accumulation after a relatively quiescent prepubertal period when mutation burdens may be plateaued.

## DISCUSSION

Here we show that lower sex- and parental-age-adjusted germline mutation rates in young adults predict longer lifespans for those same individuals, and higher parity and older age at last birth for women. Therefore, germline mutation accumulation rates in young adults may provide a measure, at least in part, of the rates of both reproductive and systemic aging. To our knowledge this is the youngest age range yet in which a molecular biomarker measured in healthy individuals has been found to predict remaining life expectancy. We encourage replications of our analyses in other populations, especially those with counts of germline mutations and clinical follow-up data already in hand, to determine whether our findings and conclusions will be corroborated.

While mutation accumulation rates are much lower in germline than in soma,^26^ many of the effectors of DNA damage and the repair systems defending against it are shared across tissues.^1,2,10,32,33^ Therefore, ranking sex- and age-matched individuals by their germline mutation accumulation rates may effectively also rank them by their somatic mutation accumulation rates, in which case the associations we have identified may be interpreted as providing strong support for the somatic mutation theory of aging.

Several recent studies^34-36^ have reported that somatic mutations in blood nuclear DNA can be detected and quantified in nearly all healthy middle-aged and older individuals, and that higher somatic mutation levels predict higher all-cause mortality. However, there is little data on the dynamics of accumulation of somatic mutations in non-tumor tissues at younger ages. At present most of the relevant available data come from analyzing DNA sequences from tumors, which include somatically acquired neutral (i.e. not tumor-promoting) mutations that were present in the cell of origin prior to tumorigenesis. By sequencing DNA from many tumors that developed at various ages across the lifespan, it has been shown that somatic mutation accumulation begins in embryogenesis, pauses from approximately ages 5 to 14 years, and then doubles thereafter approximately every eight years.^37,38^ Further work is needed to determine whether this pattern also holds true for somatic mutations in non-tumor tissue. Remarkably, intrinsic mortality rates are also plateaued from approximately age 5-14 years, and double every eight years thereafter;^30^ and the loss of health due to acquiring any chronic morbidity begins in the teenage years and also doubles every eight years.^39^ Thus somatic mutation accumulation after puberty may be a molecular correlate and major underlying determinant of both the increasing risk of death with increasing age and the duration of the human healthspan.

We also find that cross-sectional and longitudinal germline mutation accumulation data, taken together, suggest a model (Fig. S4) whereby there is a plateau of mutation counts prepubertally, followed by a puberty-induced resumption of mutation accumulation, at rates that vary more than 3-fold between individuals. Importantly, polygenic risk scores for later onset of puberty are associated with longer lifespans in both sexes;^40^ and later puberty is associated with decreased all-cause mortality in women,^41,42^ later menopause,^43^ and reduced risk of cancer in both sexes;^44^ as would be expected if puberty triggers the resumption of mutation accumulation, and consequently aging, in both germline and somatic tissues.

After puberty, both metabolic rate, which correlates with DNA damage rates, and DNA repair genes’ expression levels decline with age^45,10^ and their rates and relative levels of decline are likely to vary between individuals, due to both heritable genetic factors and differences in environmental exposures, including diet, exercise, and other lifestyle choices. This range of influences, many modifiable by personal choice, likely produces substantial inter-individual variation in both germline and somatic mutation accumulation rates and, therefore, rates of aging. However, significant variation in healthspan and lifespan may be expected even among individuals with identical puberty timing and mutation accumulation rates, if mutations are randomly distributed across the genome, only occasionally having pathogenic consequences.

While investigations of the causes of variation in the rate of aging in adult populations are likely to lead to novel therapies to postpone frailty and extend the human healthspan, further study of the effects of puberty on mutation accumulation rates may also lead to important medical breakthroughs. Direct measurements of somatic mutation levels in normal (non-tumor) tissues from healthy subjects throughout childhood and adolescence are needed to either build support for or refute the hypothesis that the mutation levels are plateaued prepubertally. Rather than always rising with age, mortality rates are known to be plateaued in some contexts, e.g. in humans during the prepubertal years^30^ and at age 105 or older,^46^ and in hydra^47,48^ and asexual planaria^49^ throughout life. These mortality plateaus may share gene expression profiles that robustly maintain the integrity of the genome (or at least prevent its further deterioration) and maintain other aspects of homeostasis as well,^3^ effectively putting aging on hold.

Interventions in adults directed toward returning mutation accumulation rates to the negligible or very low levels that may be present prepubertally would be expected to have broad benefits, greatly lowering the risks for multiple aging-related diseases and dramatically extending the human healthspan. Perhaps a relatively small number of genes that are master regulators of gene networks maintaining genome stability and homeostasis generally are downregulated at puberty, but can be reprogrammed^50^ or otherwise coaxed back to their prepubertal levels of activity by a combination of lifestyle, dietary, and/or pharmacological interventions.

## Contributors

RMC conceived the idea for the study and wrote the original draft. MFL and others ascertained and enrolled the Utah CEPH families to build the first comprehensive human genetic linkage map, and administered the UGRP. LB curated the Utah CEPH and UGRP records and DNA samples. MMD served as Study Coordinator for the UGRP. APP served as the Medical Director for the UGRP. LBJ and others acquired the funding and administered the project to obtain full genome sequences for the research subjects. TAS and ARQ curated the sequence data, identified and validated the *de novo* mutations, and assigned them to the Generation I subjects and Generation II parental couples in whom they originated. HDM, KRS, RAK, and EO performed all statistical tests of the association of germline mutation rates with the aging-related phenotypes provided by the UPDB and the UGRP. All authors provided critical feedback and helped to shape the research and data analysis, and the review and editing of the manuscript. HDM and TAS contributed equally to this study.

## Data Availability

Most of the data analyzed in this manuscript are provided either within the manuscript itself, or in the manuscript posted by Sasani et al. on bioRxiv at https://www.biorxiv.org/content/10.1101/552117v2 and its accompanying links; additional data may be accessed by contacting the corresponding author (Dr. Cawthon).

## Declaration of interests

The authors declare that they have no potential conflicts of interest relevant to this study.

## Acknowledgments

We thank all the Utah individuals who participated in the CEPH consortium and all family members who participated in the UGRP. We thank Ray White, Ph.D. (deceased), and Jean-Marc Lalouel D.Sc., for their leadership in ascertaining and enrolling the Utah CEPH families in the 1980s to build the first comprehensive human genetic linkage map; and Stephen M. Prescott, M.D. for his leadership in envisioning and building the UGRP. We thank Alison M. Fraser, MSPH for conducting many queries of the UPDB. We thank Brent S. Pedersen, Ph.D. and Jeff Stevens for helpful discussions.

## Appendix

### I. Supplemental Tables

**Table S1:** Demographic Characteristics of Utah CEPH Grandparents (n = 122)

|  | Men (N = 61) | Women (N = 61) |
| --- | --- | --- |
| Number of germline autosomal mutations | 51.1 ± 11.9 | 12.9 ± 4.3 |
| Germline mutation rate* | (9.9 ± 2.3) × 10<sup>-9</sup> | (2.5 ± 0.8) × 10<sup>-9</sup> |
| Birth year | 1909.7 ± 7.7 | 1912.6 ± 7.2 |
| Born in Utah |  |  |
| - Yes | 39 (63.9%) | 36 (59.0%) |
| - No | 18 (29.5%) | 24 (39.3%) |
| - Unknown | 4 (6.6%) | 1 (1.6%) |
| White |  |  |
| - Yes | 53 (86.9%) | 56 (91.8%) |
| - No | 0 (0.0%) | 0 (0.0%) |
| - Unknown | 8 (13.1%) | 5 (8.2%) |
| Hispanic |  |  |
| - No | 48 (78.7%) | 50 (82.0%) |
| - Yes | 1 (1.6%) | 2 (3.3%) |
| - Unknown | 12 (19.7%) | 9 (14.8%) |
| Age at index births (Parental Age) | 29.1 ± 5.6 | 26.3 ± 5.2 |
| Number of live births | 6.0 ± 2.6 | 6.0 ± 2.6 |
| Age at last birth | 39.9 ± 6.8 | 37.0 ± 6.3 |
| All births had birth dates |  |  |
| - Yes | 51 (83.6%) | 51 (83.6%) |
| - No | 10 (16.4%) | 10 (16.4%) |
| Age at menarche (n = 20) | - | 12.8 ± 0.9 |
| Cancer diagnosis |  |  |
| - No | 43 (70.5%) | 45 (73.8%) |
| - Yes | 18 (29.5%) | 16 (26.2%) |
| Age at first cancer diagnosis | 76.4 ± 8.9 | 75.6 ± 11.3 |
| Death year | 1995.8 ± 8.0 | 2000.6 ± 9.7 |
| Died in Utah |  |  |
| - Yes | 46 (75.4%) | 48 (78.7%) |
| - No | 5 (8.2%) | 9 (14.8%) |
| - Unknown | 10 (16.4%) | 4 (6.6%) |
| Censor in 2018 |  |  |
| - Alive | 1 (1.6%) | 1 (1.6%) |
| - Death | 60 (98.4%) | 60 (98.4%) |
| • Infection | 0 (0.0%) | 1 (1.6%) |
| • Cancer | 4 (6.6%) | 4 (6.6%) |
| • Endocrine/Metabolic | 2 (3.3%) | 4 (6.6%) |
| • Mental/Psychoneurotic | 1 (1.6%) | 2 (3.3%) |
| • Nervous system | 1 (1.6%) | 2 (3.3%) |
| • Circulatory system | 30 (49.2%) | 24 (39.3%) |
| • Respiratory | 4 (6.6%) | 2 (3.3%) |
| • Digestive System | 1 (1.6%) | 2 (3.3%) |
| • Other | 3 (4.9%) | 6 (9.8%) |
| • Unknown | 14 (23.0%) | 13 (21.3%) |
| *(#germline autosomal mutations/#diploid autosomal callable basepairs) |  |  |
| **Table S1: Demographic Characteristics of Utah CEPH Grandparents (n = 122)** |  |  |

**Table S2:** Median age of subjects in each category of age-adjusted mutation rates.

|  | Quartiles |  |  |  | Tertiles |  |  |
| --- | --- | --- | --- | --- | --- | --- | --- |
|  | < 25<sup>th</sup><br>percentile | 25<sup>th</sup>-50<sup>th</sup><br>percentile | >50<sup>th</sup>-75<sup>th</sup><br>percentile | > 75<sup>th</sup><br>percentile | < 33<sup>rd</sup><br>percentile | 33<sup>rd</sup> – 66<sup>th</sup><br>percentile | > 66<sup>th</sup><br>percentile |
| **Males** | 27.5 | 30 | 27.7 | 28.3 | 27.4 | 29 | 27.7 |
| **Females** | 25.9 | 25.6 | 24.5 | 26.7 | 25.9 | 25.1 | 26.6 |

**Table S3:** Effect of [germline mutation rate / parental age] on mortality in 122 Generation I individuals.

| Age-adjusted<br>germline<br>mutation rates | Both sexes |  | Males |  | Females |  |
| --- | --- | --- | --- | --- | --- | --- |
|  | HR (95% CI) | p | HR (95% CI) | p | HR (95% CI) | p |
| Continuous | 1.97 (1.12, 3.46) | **0.019** | 1.54 (1.05, 2.25) | **0.025** | 1.26 (0.92, 1.72) | 0.158 |
| 25<sup>th</sup>-50<sup>th</sup> percentile | 1.28 (0.75, 2.19) | 0.359 | 0.91 (0.43, 1.95) | 0.816 | 2.10 (0.94, 4.67) | 0.069 |
| >50<sup>th</sup>-75<sup>th</sup> percentile | 1.61 (0.95, 2.73) | 0.080 | 1.74 (0.83, 3.65) | 0.140 | 1.42 (0.64, 3.15) | 0.392 |
| > 75<sup>th</sup> percentile | 1.73 (1.02, 2.93) | **0.042** | 1.84 (0.84, 4.05) | 0.130 | 1.81 (0.85, 3.89) | 0.125 |
| Trend test | 1.20 (1.02, 1.42) | **0.028** | 1.29 (1.00, 1.66) | **0.048** | 1.15 (0.91, 1.45) | 0.235 |
| Hazard ratios (HR) and 95% Confidence Intervals (CI) were estimated using Cox proportional hazard models to assess the association of germline mutation rates per parental age [(#autosomal mutations / diploid autosomal callable basepairs) / (Parental Age)] as a continuous variable and as quartiles with all-cause mortality risk of CEPH Generation I subjects and additionally adjusted for birth year. Time was measured in years from Parental Age to time of death or last known living dates (as of 2018). Analyses were performed for both sexes combined and for each sex separately. The first row (Continuous) presents the impact on the Hazard Ratio (HR) of a one standard deviation increase in age-normalized mutation rates. The second, third, and fourth rows present the mortality risks to subjects with increasing quartiles of age-normalized mutation rates, expressed relative to the mortality risks for the lowest quartile (<25<sup>th</sup> percentile). |  |  |  |  |  |  |

**Table S4:** Effect of age-adjusted germline mutation rates on cancer risk.

| Age-adjusted<br>germline<br>mutation rate | Both sexes |  | Males |  | Females |  |
| --- | --- | --- | --- | --- | --- | --- |
|  | HR (95% CI) | p | HR (95% CI) | p | HR (95% CI) | p |
| Continuous* | 1.19 (0.83, 1.69) | 0.340 | 1.47 (0.87, 2.48) | 0.149 | 0.74 (0.44, 1.24) | 0.259 |
| 33<sup>rd</sup> – 66<sup>th</sup> percentile | 1.03 (0.42, 2.50) | 0.952 | 0.70 (0.16, 2.97) | 0.626 | 1.08 (0.31, 3.74) | 0.897 |
| > 66<sup>th</sup> percentile | 1.50 (0.66, 3.41) | 0.334 | 2.53 (0.85, 7.50) | 0.094 | 0.60 (0.15, 2.37) | 0.463 |
| Trend test | 1.24 (0.81, 1.88) | 0.318 | 1.73 (0.96, 3.12) | 0.071 | 0.77 (0.40, 1.48) | 0.440 |
| Hazard ratios were estimated using Cox proportional hazard models to assess the effect of AAMRs as a continuous or categorical variable on cancer risk of CEPH Generation I subjects, and additionally adjusted for birth year and parental age. Time was measured in years from parental age to time of death or last known living dates (up to 2018) or first cancer diagnosis, whichever occurred first. The first row (Continuous) presents the effects on cancer risk of a one standard deviation increase in AAMRs. The next two rows present the cancer risks for subjects in the middle and top tertiles for AAMRs relative to subjects in the bottom tertile. (Tertiles rather than quartiles were analyzed to provide more stable risk estimates, given the small number of cancer cases.) The cut points for AAMRs are: Males: 33% = - 0.9104878, 66% = 0.7600077; Females: 33% = - 0.34799546, 66% = 0.30450212. HR (95% CI): Hazard Ratio and 95% Confidence Interval. |  |  |  |  |  |  |
| **Table S4: Effect of age-adjusted germline mutation rates on cancer risk** |  |  |  |  |  |  |

### II. Supplemental Figures

**Figure S1.**
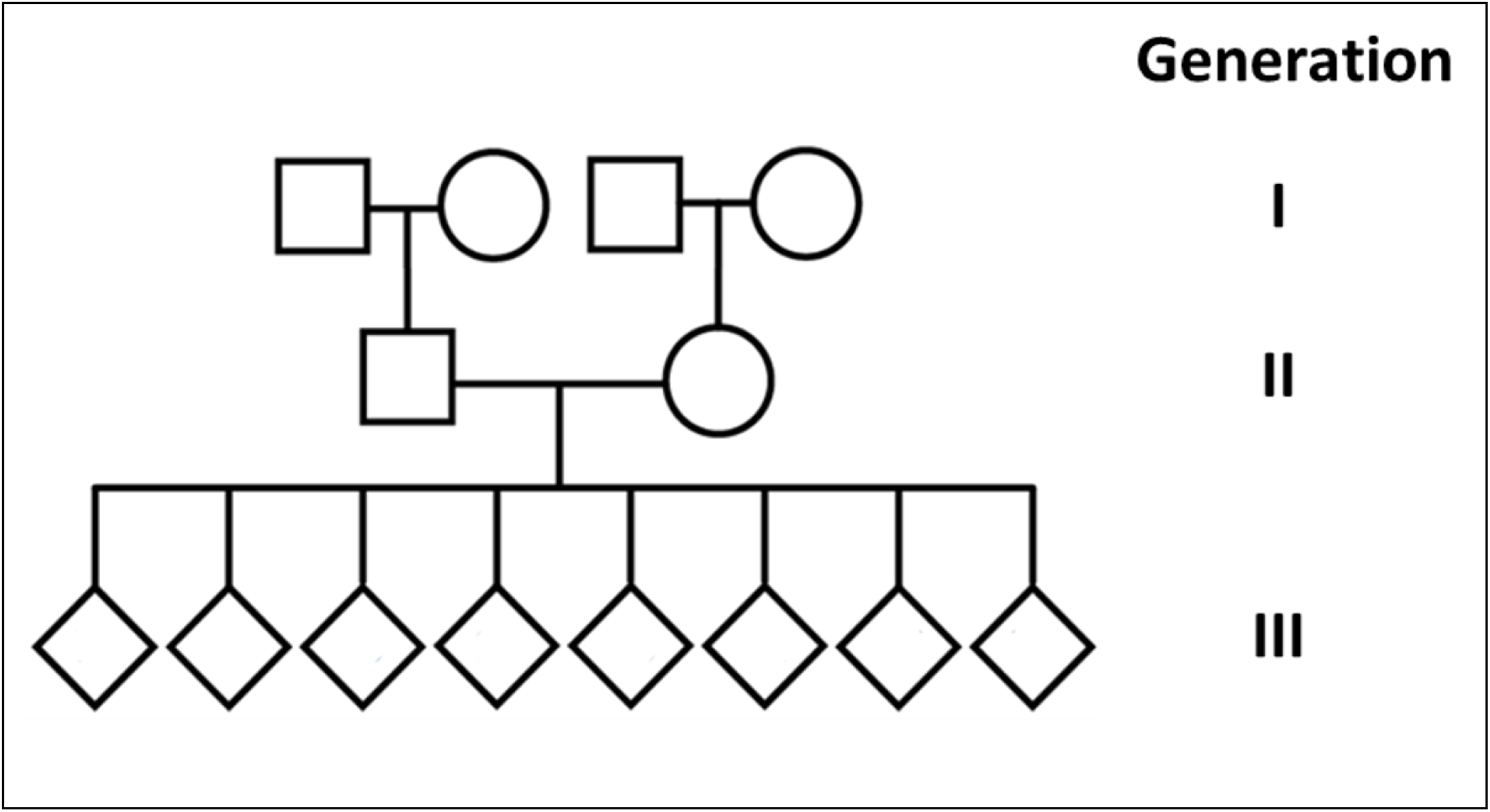
Pedigree structure of Utah CEPH three-generation families. Circles denote females, squares males, and diamonds third-generation grandchildren of either sex. Generation III sibship size ranged from 4-16. DNA sequence variants discovered in the blood DNA of Generation II individuals had to be absent from the blood DNA of their Generation I parents and present in one or more of their Generation III offspring to be considered candidates for *de novo* mutations that originated in Generation I germ cells. Additional criteria were applied to exclude from consideration early post-zygotic mutations in Generation II individuals (Sasani et al.^19^, pp. 9-10).

**Figure S2.**
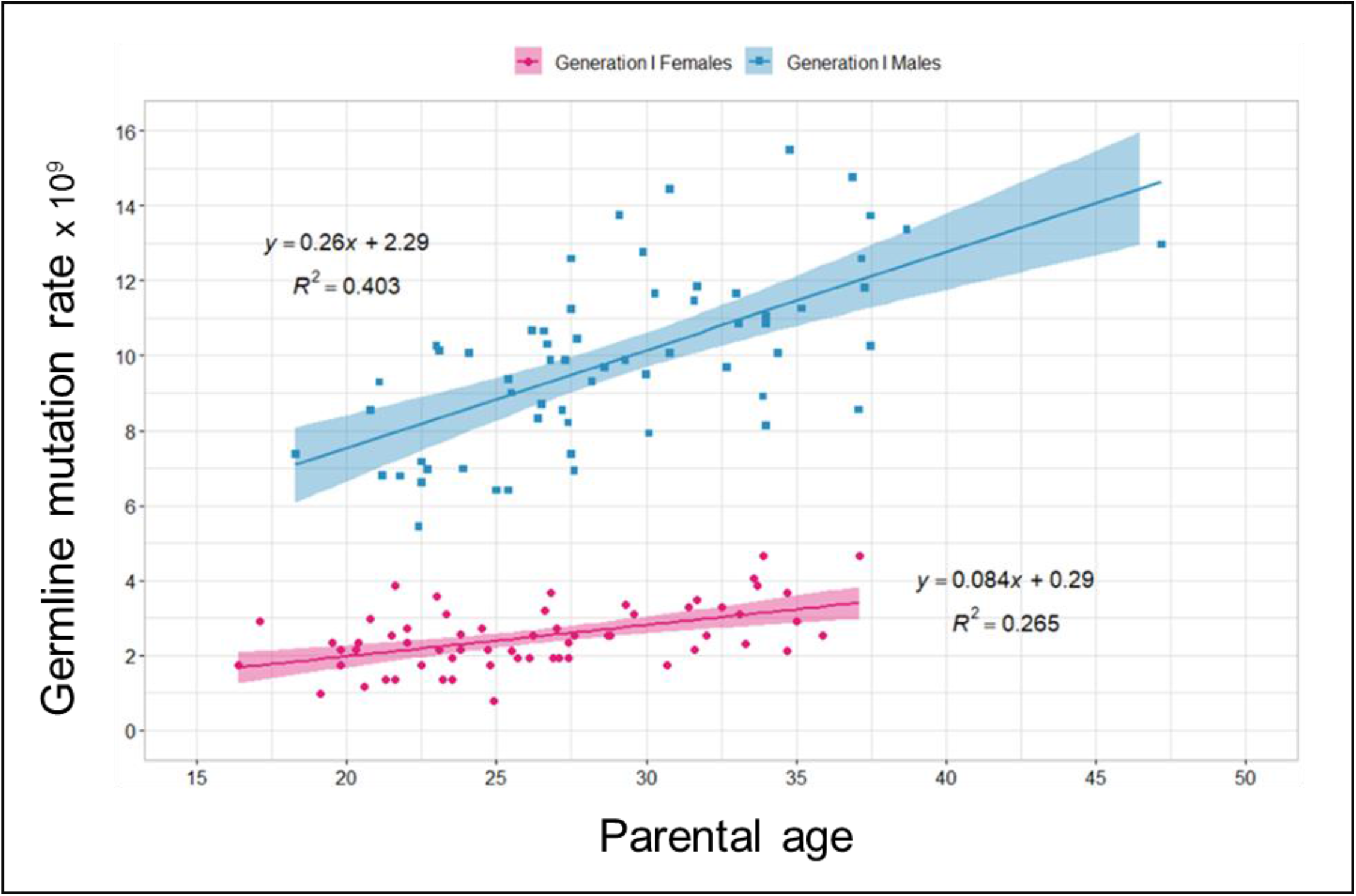
Increasing numbers of mutations in Generation I germ cells with increasing age. (Adapted from Sasani et al.^19^, Fig. 2a.) The linear regression lines and equations best fitting the data are shown. The single datapoint plotted for each of the Generation I subjects (61 men and 61 women) is derived from *de novo* mutations discovered in a single offspring. Note for each sex the more than two-fold range of mutation rates found in approximately age-matched individuals. Differences between Generation I individuals in their germline mutation rates are unlikely to be due to differences across the cohort in the presence or absence or degree of progression of various terminal illnesses, since all Generation I subjects survived more than 20 years past the age at which they transmitted these germ cell mutations to their offspring. Furthermore, it is unlikely that any of the mutations analyzed here are strongly deleterious, since all Generation II individuals in whom the *de novo* mutations were identified are known to have reached maturity and had several children of their own. Germline mutation rate = (#germline autosomal mutations) / (#diploid autosomal callable basepairs).

**Figure S3.**
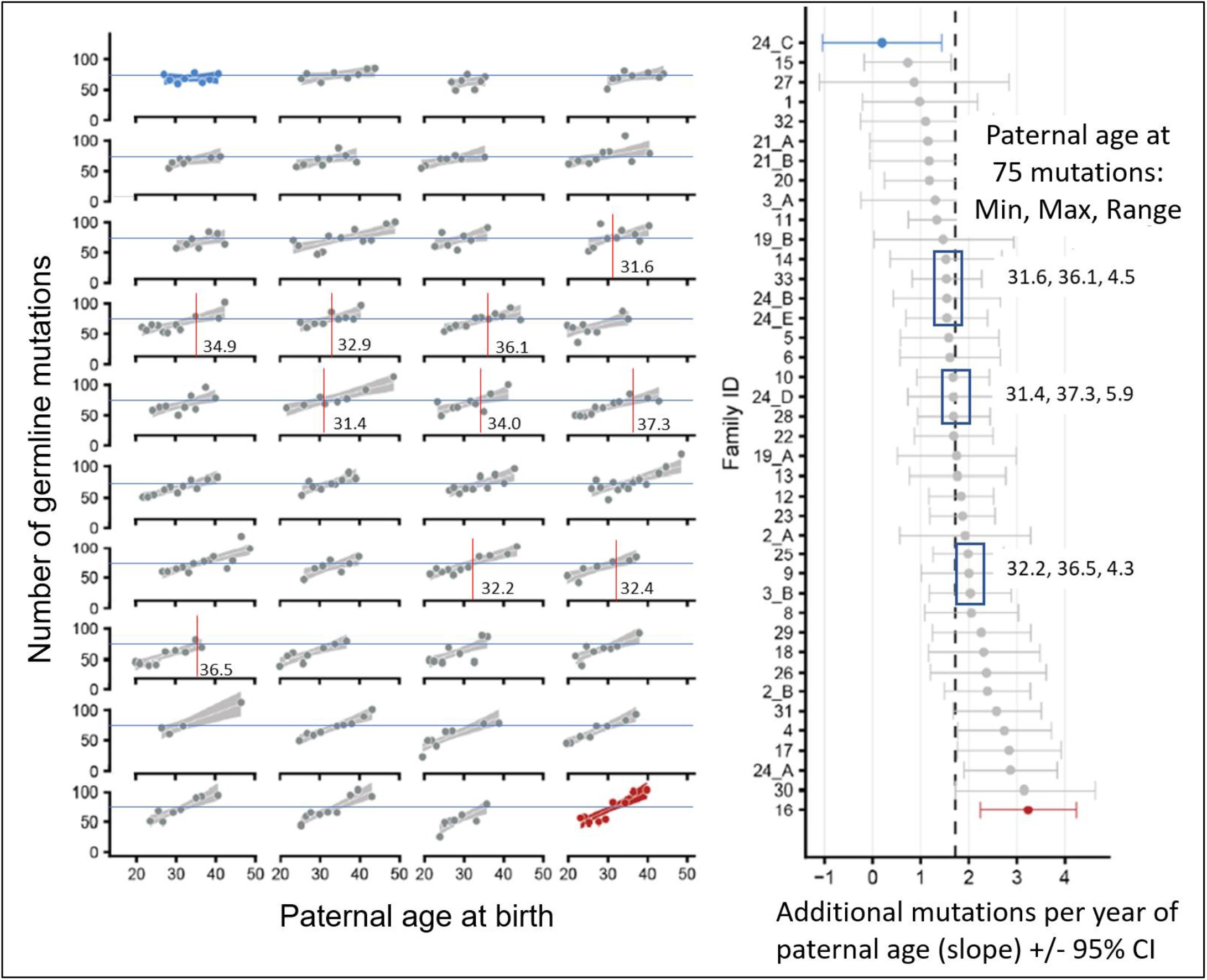
Generation II parental couples with similar germline autosomal mutation accumulation rates vary ∼5 years in the paternal age at which equivalent numbers of mutations are accumulated. Adapted from reference 19, Fig. 3c and 3d. Left panel: longitudinal data for *de novo* mutations (DNMs) discovered in 40 Generation III sibships, attributable to the germlines of the corresponding 40 Generation II parental couples. Each of the 40 plots shows the numbers of DNMs found in each sibling in a single sibship. The plots are presented, left to right and top to bottom, in order of increasing germline mutation accumulation rates. The mutation counts mainly reflect the fathers’ mutations, as the fathers have higher baseline levels and higher mutation accumulation rates than the mothers. In each row the horizontal blue line corresponds to 75 mutations accumulated, and the vertical red lines identify the paternal age at which the 75 mutations are projected to have accumulated. These paternal ages, derived from equations fitting linear regression lines to each sibship’s datapoints, are written to the right of each vertical red line. Right panel: the mutation accumulation rates for the 40 couples are listed from top to bottom in order of increasing rate. Note the more than 3-fold range of mutation accumulation rates. Blue boxes identify subsets of parental couples with very similar within-subset rates. To the right of each blue box, the minimum, maximum, and range for the paternal age needed to accumulate 75 mutations is given. The average for this range, across the three subsets, is 4.9 years, suggesting that the age at which adult germline mutation accumulation rates become established varies approximately five years, which is also the reported range for the age of onset of puberty.^31^

**Figure S4.**
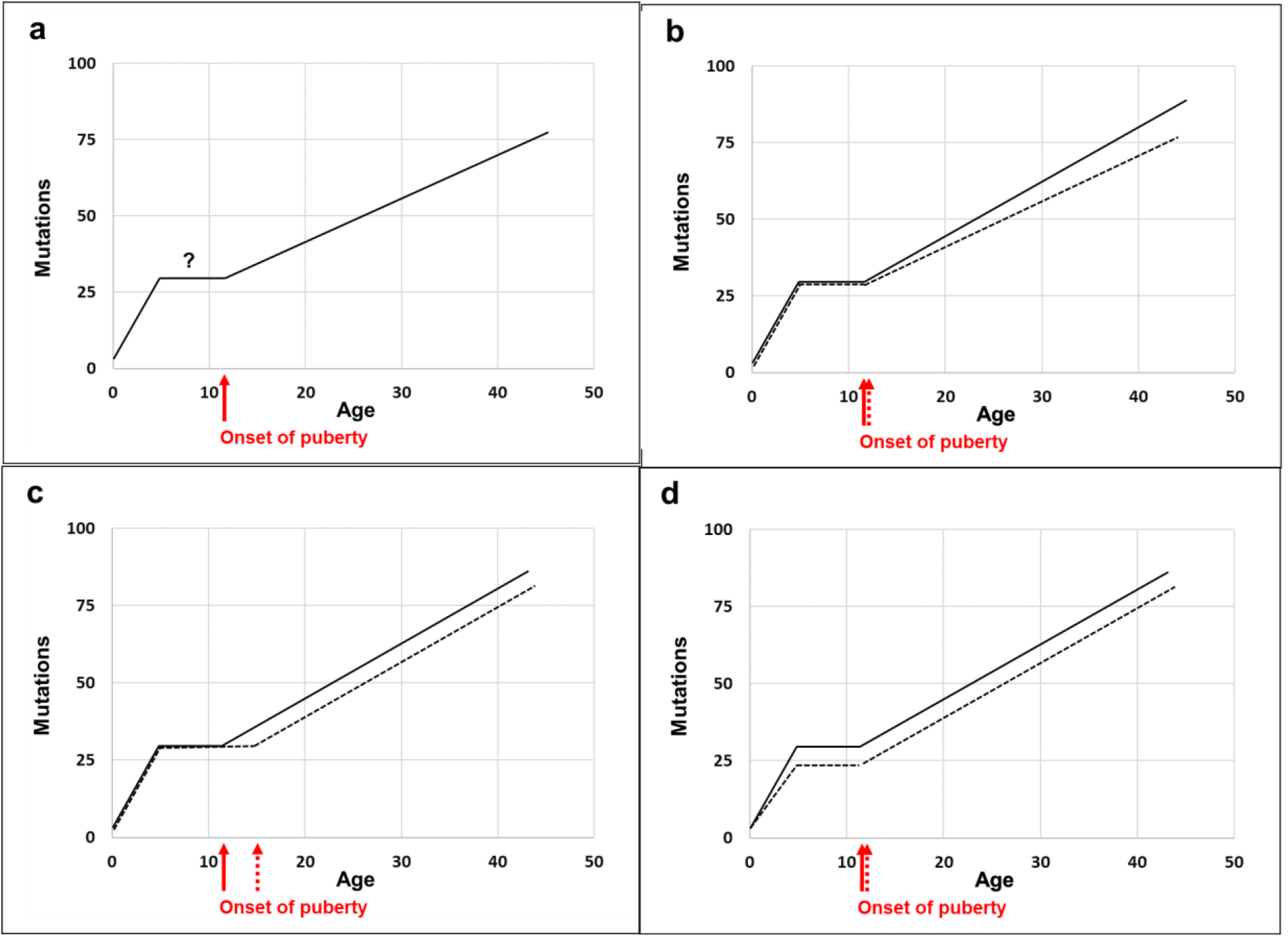
Model for germline mutation accumulation with age, with likely sources for inter-individual variation in age-specific mutation levels. (a) Rapid mutation accumulation during early childhood, followed by prepubertal plateau, followed by puberty-induced resumption of mutation accumulation. (b) Differences in mutation accumulation rates in two individuals with the same age of onset of puberty. (c) Differences in age of onset of puberty in two individuals with the same mutation accumulation rate. (d) Differences in the level of the prepubertal plateau. (Male germline mutation rates are modeled here; the same principles apply to the modeling of female germline mutation rates.)

## Notes

### Competing Interest Statement

The authors have declared no competing interest.

### Author Declarations

All relevant ethical guidelines have been followed and any necessary IRB and/or ethics committee approvals have been obtained.

Any clinical trials involved have been registered with an ICMJE-approved registry such as ClinicalTrials.gov and the trial ID is included in the manuscript.

